# Cost effective reusable gowns for routine use in hospital and laboratory: a necessity arising of COVID-19 Pandemic

**DOI:** 10.1101/2020.06.11.20127993

**Authors:** Priyanka Shahane-Kapse, Anjali Patond, Vijayshri Deotale, Rahul Narang

**Affiliations:** Department of Microbiology, Mahatma Gandhi Institute of Medical Sciences, Sevagram, Wardha, India

## Abstract

COVID-19 pandemic caused by SARS-CoV-2 has started a paradigm shift in working in a hospital or laboratory and airborne precautions have become equally significant as universal blood and body fluid precautions. Use of PPE including surgical masks or N95FFR is becoming a norm and this has led to scarcity of PPE for healthcare workers. We have prepared gowns for healthcare and laboratory workers made from a reusable material and tested with various disinfectants and heat. The cloth could tolerate repeated exposures to heat, alcohol, hypochlorite and hydrogen peroxide, but was damaged by phenol. Since this impermeable material did not allow the air to pass, we used a cool vest made up of indigenous “Khadi” cloth with pockets containing phase change material. The cost of whole reusable assembly of gown and cap was Indian Rupees 250 (1USD=76INR). This can be used in healthcare workers in hospital and community as well as people in diagnostic and research laboratories as a cost effective PPE.

## Introduction

SARS Coronavirus-2 (SARS-CoV-2) the agent that causes COVID-19 is a positive sense single stranded RNA virus belonging to subgenus Sarbecovirus of genus Betacoronavirus.^1^ The disease started in China in December 2019 and has now become a pandemic. According to recent figures a total of 7.46 million people across globe have suffered from COVID-19 with 419,000 deaths.^2^ This disease has fast emerged in India and other developing countries primarily in major metropolitan cities, but slowly spreading to smaller towns and villages as the population started migrating to their homes from cities. It is a well known fact that patients presenting with any other clinical illness may carry SARS-CoV-2 in pre clinical or asymptomatic stage^3^ and the routinely received respiratory and intestinal samples of such subjects may carry the virus along with other pathogen^4,5^. This may be universally applicable to all the patients in a hospital as we do not know the COVID-19 status of a subject unless she is tested for the same. Even, the most sensitive test, RT PCR detects only 29-93% of those carrying SARS-CoV-2 in various samples.^6^ As the virus is highly infectious and may not induce the immune-protective response^7^, the chances of the virus to be there in population will be high as endemic strain in the future. But till this happens, a lot of precautions are warranted for community as well as healthcare workers.

The route of transmission of SARS-CoV-2 is not ascertained and an April 2, 2020, expert consultation from the National Academies of Sciences, Engineering, and Medicine to the White House Office of Science and Technology Policy concluded that available studies are consistent with the potential aerosol spread of severe acute respiratory syndrome coronavirus 2 (SARS-CoV-2), not only through coughing and sneezing, but by normal breathing.^8^ Diagnosis of COVID-19 has led to revamping of various laboratories and a number laboratories have been repurposed for COVID-19 testing. In India, Board of Governors of the erstwhile Medical Council of India have released an order recently that all the medical colleges should have a biosafety level-2 (BSL-2) laboratory for infectious pathogens in the department of Microbiology. Another report from Nature has indicated that as scientists around the world return to work, they’re encountering new safety rules and awkward restrictions — and sometimes writing the protocols themselves^9^.

This paradigm shift warrants that the healthcare workers and researchers become more inclined to airborne safety in addition to universal safety precautions.

Similar to COVID-19, cough is a common feature of tuberculosis (TB) and several other diseases and precautions are most needed for laboratory diagnosis of TB. It has been recommended that Biosafety Level-1 without a biosafety cabinet may be sufficient to handle sputum samples for TB smear microscopy and cartridge based nucleic acid amplification test (CB NAAT). For COVID-19 testing the laboratories should have at least Class 2 A2 or B biosafety cabinets in BSL-2 laboratory. This has to become a norm for all the samples as the patients excrete SARS-CoV-2 in various samples^5^.

As the pandemic progresses there has been shortage of PPE including N95FFR. Recently, we conducted a study to analyse effect of various disinfecting techniques for respirators and prepared SOP for the same.^10^ Since disposable laboratory gowns are becoming scarce, we are proposing use of reusable in-house gowns for the laboratory as well as healthcare workers.

## Material and methods

To prepare and test an in-house reusable gown we contacted Defense Research and Development Organization (DRDO) as they had been working on selection of such material for their own PPE. (Personal Communication) After discussion, we contacted a company (AjyTech, Surat India https://www.ajytechindia.com/) and they agreed to provide us cloth for testing and preparation of at least 2 gowns. Before, this cloth could be used we tested its efficacy, integrity, durability, resistance to various decontamination procedures and acceptability by the end users. The vendor had claimed that this cloth, made up of polyester coated with polyurethane could be reused at least 20 times after washing.

For various tests, we made 15×15cm square pieces of the cloth.

Testing for impermeability: This was tested by making a pocket of the cloth and holding 5ml of water for 5 minutes. (photo 1)

**Photo 1:**
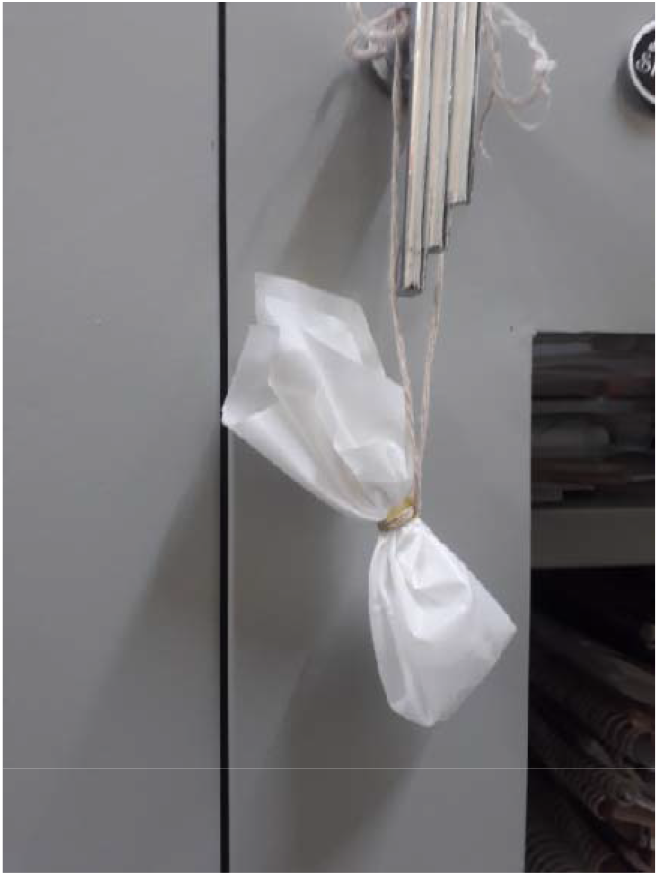
Water retention capacity.

Testing for various decontamination procedures: Various pieces of this cloth were subjected to repeated exposure to 70% ethanol, 5% phenol, 1% hypochlorite, 1% H2O2, boiling at 100deg C and autoclaving at 121 deg C for 15min each.

For checking its integrity and durability-After every treatment, we repeated the water holding experiment. Gross examination of the cloth was conducted every time any experiment was performed.

Acceptability by end users: The cloth was shared with the laboratory people and based on their choice we got it stitched by our hospital linen section in line with how they stitch surgical gowns for operation theater.

## Results

The cloth passed the water holding technique after treatment with all the disinfectants and heat including autoclaving, except Phenol treatment where the material was damaged. (Photo 2) Physically, there was no change in the cloth except for some insignificant creases after repeated 70% alcohol treatment. The laboratory staff liked the cloth as it was lighter than other disposable PPE they were using. However, they had an apprehension that since water could not pass through it air will also not pass. For this purpose, after brain storming session we decided to make it in the form of gown to allow air to pass from below and use it wherever air-conditioning was available in the laboratory.

**Photo 2:**
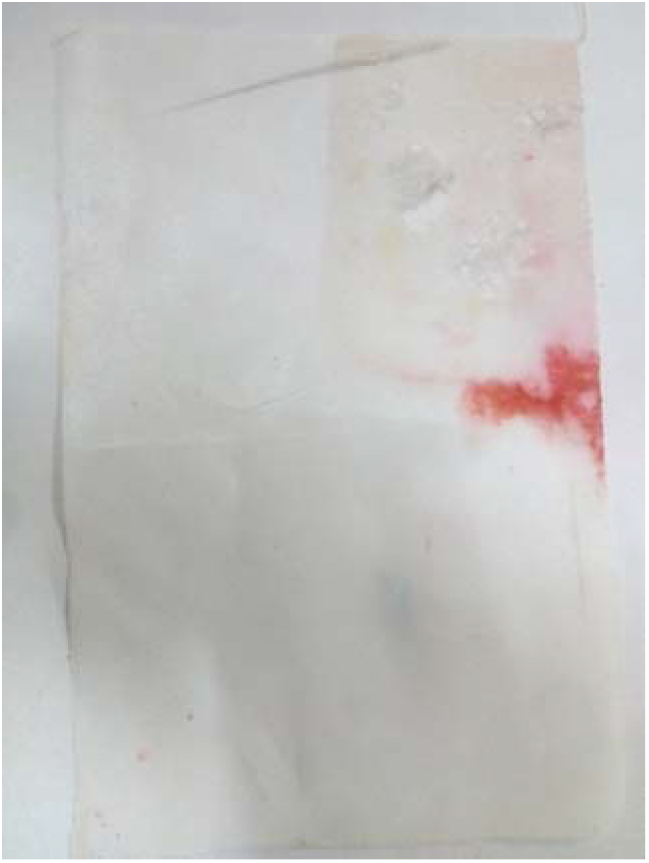
Cloth damaged after phenol treatment experiment.

Also, at the request of laboratory staff, we improvised a “Khadi” (homespun cotton) vest with four large pockets to accommodate pouches of phase change material^11^ that was kept in refrigerator before use. “Khadi” cloth by virtue of its make absorbs sweat in warm humid climate and is well accepted in our area with such conditions.

The hospital tailor was able to stitch it into a gown by using 2.5 m of cloth. The cost for one gown was just Rs 250 at the rate of Rs 100 per meter. (1USD=76 INR)

## Discussion

The COVID-19 pandemic has started affecting the developing countries and the hospitals and laboratories have started struggling for procurement of PPE.

Hospitals have started reusing the single use PPE by using various techniques, e.g. Hypochlorite, H2O2, Ethylene Oxide or a combination of these. The material used for reusable PPE is not sturdy and after few uses starts to disintegrate. The material tested in the present study has been shown to sustain repeated use of disinfectants, except phenol, and heat for at least 20 times. Any of the methods can be used for disinfection of the gown. One precaution is for the use of hypochlorite as disinfectant; where after treatment the gown has to be properly washed with water otherwise it becomes stained and sticky. (Photo 3)

**Photo 3:**
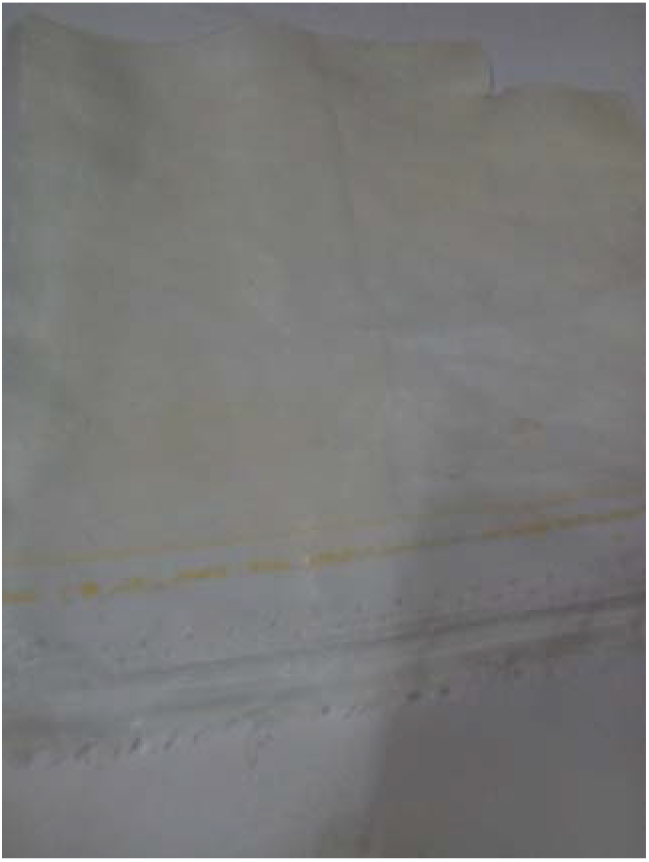
Cloth stained after repeated Hypochlorite treatment.

We also looked for the comfort of end user by stitching it in the form of a gown and using PCM inside to keep the body temperature low. JAMA had published an editorial sometime back wherein editors had asked for suggestions for the extended use or reuse of PPE^11^. One of the authors of the present study had suggested use of vests of phase change material inside the gown to protect the wearer from heat and sweat in warm and humid climate as a comment to the edotorial^11^. Since the commercially available jacket with phase change material was not available with us, we used indigenous cloth “Khadi” that solved the issue of sweating as well. The packets of phase change material were reused after keeping them in refrigerators (not freezer) and the “Khadi” clothe was washed with soap and water.

Another item for PPE include head cap that has also been made reusable using same material. In another publication, we have also suggested reuse of N95FFR using UV-C in an indigenously made cabinet made from materials available in the microbiology department.^10^ We are using reusable eye protection (Rs 300) along with other items. (Photo 4)

**Photo 4:**
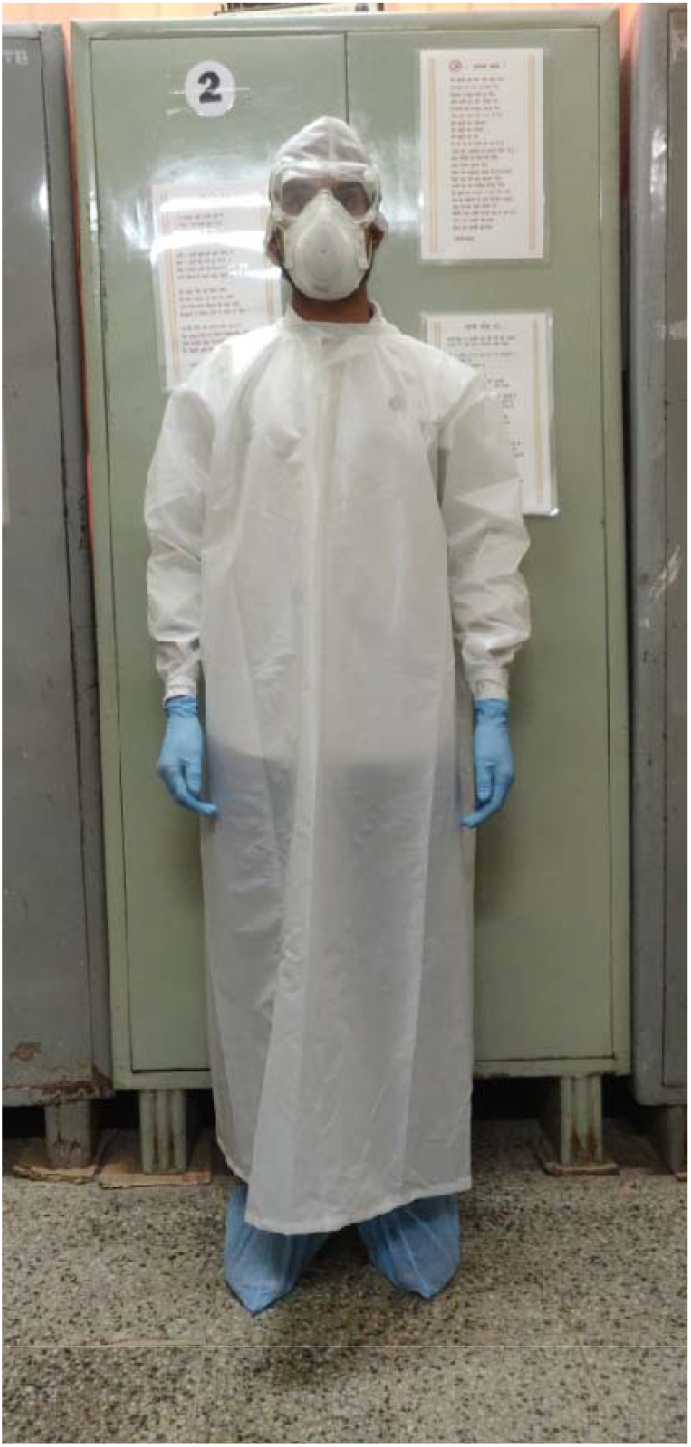
Complete cost effective reusable PPE.

## Conclusion

We conclude this cost effective reusable gown along with masks/respirators can be used in any area where protection from droplets/aerosols is required, including hospitals and laboratories not only during COVID-19 pandemic but beyond that as well.

## Data Availability

We have provided the experiment data in the form of photographs.

## Acknowledgements

We acknowledge support from Dr Rajiv Narang, DRDO for guidance, Meenakshi Shahane and Razique for photo courtesy, Linen Department of MGIMS and AjyTech, Surat.

## References

1. Zheng J. SARS-CoV-2: an emerging coronavirus that causes a global threat. Int J Biol Sci 2020;16:1678–85. http://doi.org/10.7150/ijbs.45053.

2. https://www.worldometers.info/coronavirus/ Accessed on 11.06.2020

3. Gandhi M, Yokoe DS, Havlir DV. Asymptomatic transmission, the Achilles’ heel of current strategies to control COVID-19. https://www.nejm.org/doi/full/10.1056/NEJMe2009758.

4. Pan Y, Zhang D, Yang P, Poon LL, Wang Q. Viral load of SARS-CoV-2 in clinical samples. Lancet Infect Dis 2020;20:411–2. http://doi.org/10.1016/S1473-3099(20)30113-4.

5. Lo IL, Lio CF, Cheong HH, Lei CI, Cheong TH, Zhong X, Tian Y, Sin NN. Evaluation of SARS-CoV-2 RNA shedding in clinical specimens and clinical characteristics of 10 patients with COVID-19 in Macau. Int J Biol Sci 2020;16:1698–1707. http://doi.org/10.7150/ijbs.45357.

6. Wang W, Xu Y, Gao R, Lu R, Han K, Wu G, Tan W. Detection of SARS-CoV-2 in different types of clinical specimens. JAMA. 2020 May 12;323(18):1843–4.

7. Giesecke J. The invisible pandemic. The Lancet. 2020 May 5. https://doi.org/10.1016/S0140-6736(20)31035-7.

8. National Academies of Sciences, Engineering, and Medicine. Rapid Expert Consultation on SARS-CoV-2 Laboratory Testing for the COVID-19 Pandemic (April 8, 2020). InRapid Expert Consultations on the COVID-19 Pandemic: March 14, 2020– April 8, 2020 2020 Apr 30. National Academies Press (US).

9. Subbaraman N. Return to the lab: scientists face shift work, masks and distancing as coronavirus lockdowns ease. Nature. 2020 Jun 1; 582(7810):15–6.

10. Patond A and Narang R. A low cost ingenious approach for ultraviolet decontamination of N95 filtering face-piece respirators to deal with dwindling supply during the COVID-19 pandemic. Journal of MGIMS. (Accepted for publication)

11. Bauchner H, Fontanarosa PB, Livingston EH. Conserving supply of personal protective equipment—a call for ideas. Jama. 2020 May 19; 323(19):1911-.

12. Narang R. Use of phase change material under PPE during warm season. In response to: Bauchner H, Fontanarosa PB, Livingston EH. Conserving supply of personal protective equipment—a call for ideas. Jama. 2020 May 19; 323(19):1911-.

